# Attitudes towards glaucoma genetic risk assessment in unaffected individuals

**DOI:** 10.1101/2021.10.19.21264544

**Authors:** Georgina L Hollitt, Owen M Siggs, Bronwyn Ridge, Miriam C Keane, David A Mackey, Stuart MacGregor, Alex W Hewitt, Jamie E Craig, Emmanuelle Souzeau

## Abstract

Integrating polygenic risk scores (PRS) into healthcare has the potential to stratify an individual’s risk of glaucoma across a broad population. Glaucoma is the most common cause of irreversible blindness worldwide, therefore effective screening for glaucoma endorsed by the population is highly important. This study assessed the attitude of unaffected individuals towards PRS testing for glaucoma, and sought to identify factors associated with interest in testing. We surveyed 418 unaffected individuals including those with a first-degree relative with glaucoma (n=193), those who had a recent eye examination (n=117), and general members of the community (n=108). Overall, 71.3% indicated an interest in taking a polygenic risk test for glaucoma. Interest was more likely in those who believed glaucoma to be a severe medical condition (OR 14.58, 95%CI (1.15-185.50), p=0.039), those concerned about developing glaucoma (OR 4.37, 95%CI (2.32-8.25), p<0.001), those with an intention to take appropriate measures regarding eye health (OR 2.39, 95%CI (1.16-4.95), p=0.019), and those preferring to know if considered to be at-risk or not (OR 4.52, 95%CI (2.32-8.83), p<0.001). These findings represent a valuable assessment of general public interest in glaucoma polygenic risk testing, which will be integral to the implementation and uptake of novel PRS based tests into clinical practice.

## INTRODUCTION

Glaucoma is a degenerative condition affecting the optic nerve and can result in irreversible vision loss and blindness if left untreated. Primary open-angle glaucoma (POAG) is the most common subtype and is associated with, but not dependent on, raised intraocular pressure (IOP).^1^ IOP is the major modifiable risk factor for POAG and is therefore the target of treatment approaches including topical eye drops, laser treatment, or incisional surgical intervention. Other risk factors relate to individual genetic risk, including ethnicity and family history, with a 9.2-fold increased risk for first-degree relatives of individuals with glaucoma compared to controls.^2^ Furthermore, over half of all individuals with glaucoma in developed countries and over 90% in developing countries are estimated to be undiagnosed.^3,4^ This is largely due to the asymptomatic nature of early-stage disease, along with insufficient population screening because screening is typically opportunistic. Early diagnosis is paramount given that vision cannot be restored once it is lost,^5^ and existing treatments are highly effective in preventing or slowing disease progression.^6,7^ Since glaucoma is the most common cause of irreversible vision loss in the world, improving screening methods has the potential to significantly reduce the social and economic burden of disease.

Glaucoma is one of the most heritable common complex diseases.^8,9^ Both monogenic and polygenic factors contribute to glaucoma.^8,10^ Disease-causing variants in genes such as *MYOC* and *OPTN* or copy number variants in *TBK1* account for less than 5% of POAG with Mendelian inheritance patterns.^11^ Currently, the clinical use of genetic testing for glaucoma has been limited to these genes.^11,12^

With recent advances in the scale of genome-wide association studies (GWAS), there is increasing interest in the application of polygenic risk scores (PRS) across a variety of common diseases including glaucoma. A PRS collates the combined risk of multiple common genetic risk variants into a single score, typically by weighting the relative effect size of each variant,^13^ and is commonly expressed in the form of quantiles across a population distribution. Such scores may be combined with conventional risk factors to estimate overall disease risk. PRS results represent a probability of individual disease risk and are therefore not diagnostic, and results should be interpreted in conjunction with other established clinical risk factors, in particular age.^14^

Recent studies have demonstrated the utility of glaucoma PRS in risk stratification. A recent glaucoma PRS was associated with higher glaucoma risk (top 10% PRS compared to remaining 90% glaucoma OR = 4.2) as well as more rapid disease progression, and higher treatment intensity.^15^ Individuals in the top PRS decile were at 15-fold increased risk of developing advanced glaucoma compared to the bottom decile.^15^ An IOP PRS was shown to provide further insight into the relationship between genetic variants implicated in IOP and POAG outcomes.^16,17^ Additionally, high polygenic risk confers a comparable risk to monogenic variants, despite being over 15 times more prevalent in the general population, and can also influence the penetrance and age at diagnosis.^15,18,19^ Furthermore, by stratifying individuals across the risk spectrum for developing glaucoma and likelihood of progression, unnecessary treatment which can be costly and associated with unwanted side effects may be avoided.

PRS use is largely limited to research contexts where it is used for various purposes such as testing treatment modalities and predicting treatment outcomes, testing associations between traits and/or diseases, and determining genetic overlap between disorders.^20^ However, with more data supporting the clinical validity of PRS in risk stratification, such tests may soon become part of routine clinical care. Before this can occur, it is necessary to understand how such testing may be received by the general population and what key social and behavioural elements may impact implementation. In this study, we report the attitudes of individuals without diagnosed glaucoma towards glaucoma PRS testing, and the demographic and psychosocial factors that influence this.

## METHODS

### Study Sample

This was a cross-sectional, questionnaire-based study approved by the Southern Adelaide Clinical Human Research Ethics Committee (SAC HREC) that adhered to the Revised Declaration of Helsinki. The study sample included participants from three cohorts of individuals who may be target populations for polygenic risk testing for glaucoma. The first cohort included unaffected first-degree relatives of individuals with a known glaucoma diagnosis, with participants drawn from the Australian and New Zealand Registry of Advanced Glaucoma (ANZRAG) and the Targeting At Risk Relatives of Glaucoma patients for Early diagnosis and Treatment (TARRGET) study. Participants from outpatient clinics and the community who had at least one first-degree relative with glaucoma were also included in this cohort. ANZRAG is one of the largest databases of clinical and genetic data for glaucoma in the world (regardless of glaucoma severity),^21^ while TARRGET is designed to provide educational material to first-degree relatives of individuals with glaucoma, with their personalised risk of developing the disease according to their family member’s clinical phenotype. The second cohort included people attending an optometrist for an eye assessment for conditions *other than glaucoma*, or those with no ocular health history who had undergone an eye assessment within the last six months. These participants were recruited from private (Specsavers) and public (Flinders University) optometry clinics, as well as from public hospital settings (Flinders Medical Centre and Noarlunga Hospital) in Adelaide, Australia. The third cohort comprised members of the general community without an ocular health history, who had not undergone a recent eye examination. Recruitment occurred at Flinders Medical Centre (including the Flinders Volunteer service) and Noarlunga Hospital in Adelaide, Australia and included Flinders volunteer members, patients and their relatives in outpatient hospital clinics.

### Data Collection

Socio-demographic, health, cognitive, emotional and influencing factors were used to assess association with interest in genetic testing.

#### Socio-demographic

Age, gender, ethnicity, highest level of education, and urban/rural residency were collected. Ethnicity was self-reported and classified into 10 ethnic groupings, then into categories of “European” and “non-European” ancestry. Those recorded as “unknown” were excluded. Residency was based on the Australian Bureau of Statistics census data using the participants’ postcodes. Urban residency was classified as postcodes with populations greater than 50,000 persons. Rural residency included regional, rural and remote areas of populations less than 50,000 persons.

#### Health factors

Family history, including the number of family members affected by any form of glaucoma and their degree of relation, was self-reported by participants. Eye health factors assessed included a history of myopia, most recent eye check, and the frequency of eye checks.

#### Cognitive factors

Cognitive factors were assessed through single-item measures with Likert-like scale response options. We assessed participants’ understanding of the heritability of glaucoma, perception of the severity of glaucoma and perceived likelihood of developing glaucoma.

#### Emotional factors

To assess the influence of emotion on interest in genetic testing for glaucoma, we asked participants to indicate their level of worry related to the possibility of developing glaucoma in the future.

#### Factors affecting decision to be tested and concerns

We assessed several factors which could affect participants’ decision to be tested related to their own risk, their family’s risk and advice from others. We assessed factors which would concern participants about testing, including personal anxiety, cost, future requirements and issues relating to confidentiality and implications of results. Participants could also include additional factors or comments.

#### Outcome variable

Interest in genetic testing for glaucoma was evaluated by assessing likelihood to undergo genetic testing to predict personal glaucoma risk.

#### Additional factors

Participants were asked about aspects of the test that would be considered important to know prior to undergoing genetic testing, the cost participants would be willing to pay, and their preferred method of receiving results. Participants were asked to indicate how their behaviour towards their eye health might change based on theoretical results, and the frequency of eye checks which they would be willing to undergo.

### Statistical analysis

Data were analysed using Statistics Package for the Social Sciences (Version 27.0, SPSS Inc., Chicago, USA). Descriptive statistics were used to characterise the study sample. Responses from the three groups were combined for the statistical analysis. Responses were combined into bivariate outcomes; for example ‘highly unlikely and unlikely’ were merged into an ‘uninterested’ group, and ‘likely’ and ‘highly likely’ were merged as an ‘interested’ group. Unsure responses for all questions were excluded. The association between level of interest and covariables (sociodemographic, emotional and cognitive variables) was performed using a univariate logistic regression model. Variables that had significance levels of p<0.1 in the univariate analysis were initially included in the multivariate regression model. Multivariate logistic regression models were performed to identify factors independently associated with interest in testing (p<0.05) using a backward stepwise approach. Where multiple comparisons were made on the same data, Bonferroni correction was applied.

## RESULTS

### 1. Demographic and Personal Characteristics

In total, 418 participants completed the questionnaire; 193 had at least one affected first-degree relative, 117 had had a recent eye review and 108 were from the community. In total, 243 unaffected family members in ANZRAG and TARRGET were invited to participate in the study, and 143 completed the questionnaire, yielding a response rate of 58.8%. The other 50 participants with a first-degree relative were recruited from outpatient clinics and hospital settings. The demographic and personal characteristics of each cohort and the whole study sample are shown in Table 1. In summary, 66.5% were female, 95.0% were of European ancestry, 75.4% were from an urban area, and 63.8% had an education level above secondary school. The mean age of the total cohort was 62.1 years ± 13.3 years. There was a significant difference in residency, timing of last eye check and frequency of eye checks between groups (Table 1 - significant results in bold). Participants with affected first-degree relatives, those who had a recent eye check, and members of the general community did not differ by age, gender, ethnicity and level of education (Table 1). The majority (74.9%) of participants had undergone an eye check within at least the last year and over half (55.0%) reported undergoing eye checks at least annually.

**Table 1.** Characteristics of the study sample (including individuals with a first-degree relative with glaucoma [First-degree relative], those who had undergone a recent eye check [Optometry] and general members of the community [Community])

| <u>Variable</u> | <u>First-degree relative</u><br>n = 193 | <u>Optometry</u><br>n = 117 | <u>Community</u><br>n = 108 | <u>TOTAL</u><br>n = 418 | <u>p Value</u> |
| --- | --- | --- | --- | --- | --- |
| **Age (years)**<br>Range<br>Mean (standard deviation)<br>Median<br>Missing | 33.0 - 89.8<br>61.7 (11.2)<br>62.1 | 21.0 - 89.3<br>63.2 (15.4)<br>65.4<br>n=1 | 19.4 - 94.6<br>61.5 (14.3)<br>66.3<br>n=1 | 19.4 - 94.6<br>62.1 (13.3)<br>63.3<br>n=2 | p = 0.573* |
| **Gender, n (%)**<br>Female<br>Male | 134 (69.4)<br>59 (30.6) | 73 (62.4)<br>44 (37.6) | 71 (65.7)<br>37 (34.3) | 278 (66.5)<br>140 (33.5) | p = 0.437^ |
| **Ethnicity, n (%)**<br>European ancestry<br>Non-European ancestry<br>- African<br>- Asian<br>- Hispanic<br>- Middle Eastern<br>- Mixed<br>Unknown | 185 (95.9)<br>6 (3.1)<br>2 (1.0)<br>3 (1.6)<br>0<br>0<br>1 (0.5)<br>2 (1.0) | 114 (97.4)<br>2 (1.7)<br>0<br>1 (0.8)<br>1 (0.8)<br>0<br>0<br>1 (0.9) | 98 (90.7)<br>9 (8.3)<br>0<br>3 (2.8)<br>1 (0.9)<br>2 (1.9)<br>3 (2.8)<br>1 (0.9) | 397 (95.0)<br>17 (4.1)<br>2 (0.5)<br>7 (1.7)<br>2 (0.5)<br>2 (0.5)<br>4 (1.0)<br>4 (1.0) | p = 0.028^ |
| **Residency, n (%)**<br>Urban<br>Rural<br>Unknown | 137 (71.0)<br>55 (28.5)<br>1 (0.5) | 90 (76.9)<br>21 (17.9)<br>6 (5.1) | 88 (81.5)<br>16 (14.8)<br>4 (3.7) | 315 (75.4)<br>92 (22.0)<br>11 (2.6) | p = 0.019^ |
| **Highest level of education, n (%)**<br>Primary School<br>Secondary School<br>Vocational Training<br>University<br>Unknown | 2 (1.0)<br>60 (31.1)<br>52 (26.9)<br>77 (39.9)<br>2 (1.0) | 4 (3.2)<br>43 (36.8)<br>39 (33.3)<br>31 (26.5)<br>0 | 1 (1.0)<br>39 (36.1)<br>40 (37.0)<br>28 (5.9)<br>0 | 7 (1.7)<br>142 (34.1)<br>131 (31.3)<br>136 (32.5)<br>2 (0.5) | p = 0.056^ |
| **Family History, n (%)**<br>Positive<br>Negative<br>Unknown<br>Positive (closest affected relative):<br>- First-degree<br>- Second-degree<br>- Third-degree<br>- Unknown | 193 (100.0)<br>0<br>0<br>193 (100.0)<br>0<br>0<br>0 | 9 (7.7)<br>107 (95.1)<br>1 (0.9)<br>0<br>7 (6.0)<br>1 (0.9)<br>1 (0.9) | 7 (7.4)<br>87 (80.6)<br>14 (13.0)<br>0<br>5 (4.6)<br>1 (0.9)<br>1 (0.9) | 209 (50.0)<br>209 (50.0)<br>0<br>193 (92.3)<br>12 (5.7)<br>2 (1.0)<br>2 (1.0) | p <0.001^ |
| **Last eye check, n (%)**<br>Within 6 months<br>6-12 months<br>1-2 years<br>More than 2 years<br>Never<br>Missing | 63 (32.6)<br>82 (42.5)<br>41 (21.2)<br>4 (2.1)<br>1 (0.5)<br>2 (1.0) | 117 (100.0)<br>0<br>0<br>0<br>0<br>0 | 0<br>51 (47.2)<br>30 (27.8)<br>25 (23.1)<br>0<br>2 (1.9) | 180 (43.1)<br>133 (31.8)<br>71 (17.0)<br>29 (6.9)<br>1 (0.2)<br>4 (1.0) | p <0.001^ |
| **Frequency of eye checks, n (%)**<br>3 monthly | 2 (1.0) | 1 (0.9) | 0 | 3 (0.7) | p = 0.003^ |
| 6 monthly | 9 (4.7) | 5 (4.3) | 2 (1.9) | 16 (3.8) |  |
| Annually | 107 (55.4) | 61 (52.1) | 43 (39.8) | 211 (50.5) |  |
| Every 2 years | 61 (31.6) | 32 (27.4) | 34 (31.5) | 127 (30.4) |  |
| More than every 2 years | 10 (5.2) | 13 (11.1) | 22 (20.4) | 45 (10.8) |  |
| Never | 2 (1.0) | 3 (2.6) | 6 (5.6) | 11 (2.6) |  |
| Missing | 2 (1.0) | 2 (1.7) | 1 (0.9) | 5 (1.2) |  |

\*denotes p-value calculated using one-way ANOVA. ^denotes p-value calculated using Chi-square test for Association. Differences in ethnicity were assessed between Europeans and non-Europeans ancestry

### 2. Understanding of glaucoma and perception of severity and risk

In the overall cohort, 57.7% believed glaucoma was at least somewhat hereditary, with 57.7% of those having an affected first-degree relative. A large proportion (39.5%) of the total cohort were unsure about the hereditary nature of glaucoma. The majority (91.9%) of respondents considered glaucoma to be a severe medical condition, with an approximately equivalent proportion with (47.9%) and without (52.1%) an affected first-degree relative. Perception of glaucoma as a severe condition was associated with being likely to increase the frequency of eye checks if found to be at high risk (OR 7.36, 95%CI (1.32-40.89), p=0.023). 31.8% of participants believed they were likely or highly likely to develop glaucoma in their lifetime, and 89.1% of these expressed worry about this belief. Those with at least one first-degree relative with glaucoma were more likely to believe they were at risk of developing glaucoma (OR 5.06, 95%CI (2.99-8.58), p<0.001), and were worried about this (OR 3.75, 95%CI (2.33 - 6.06), p <0.001). Being worried about the possibility of developing glaucoma was associated with a preference to know glaucoma risk (OR 2.19, 95%CI (1.40-3.43), p <0.001).

### 3. Interest in genetic risk prediction testing for glaucoma

Overall, the majority of individuals expressed an interest in genetic risk prediction testing for glaucoma, with 71.3% of respondents indicating they would be either likely or highly likely to take a test if it were available. The attitudes of each cohort are shown in Figure 1. Over half of those who were interested in testing (62.2%) also reported they would probably or definitely like to know more about glaucoma before being tested. Individuals with at least one affected first-degree relative were also more likely to be interested in genetic testing for glaucoma than those without (OR 2.90, 95% CI 1.65-5.09, p <0.001)(Table 2).

**Table 2:**
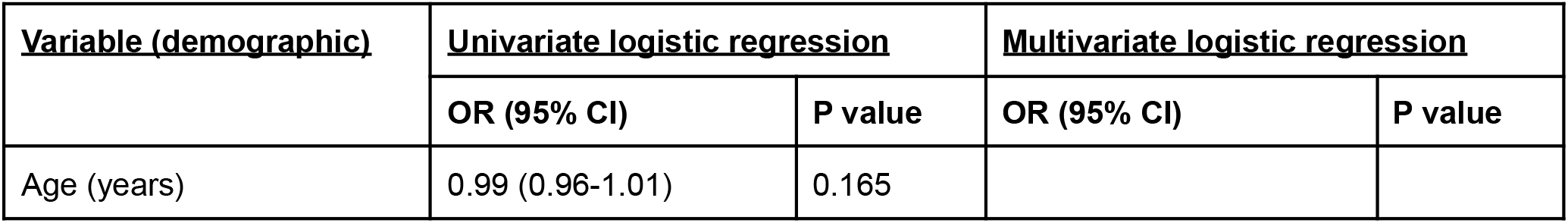
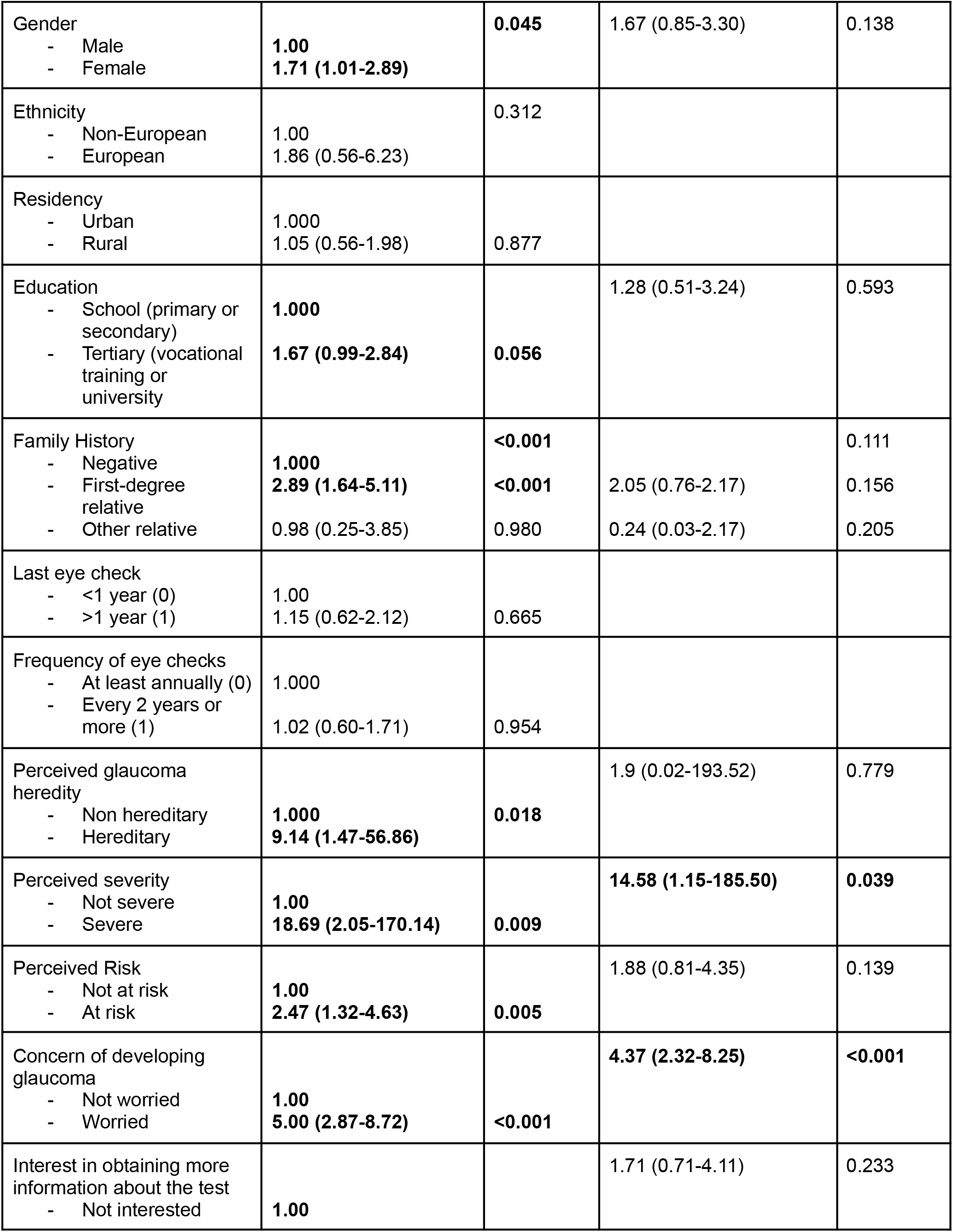
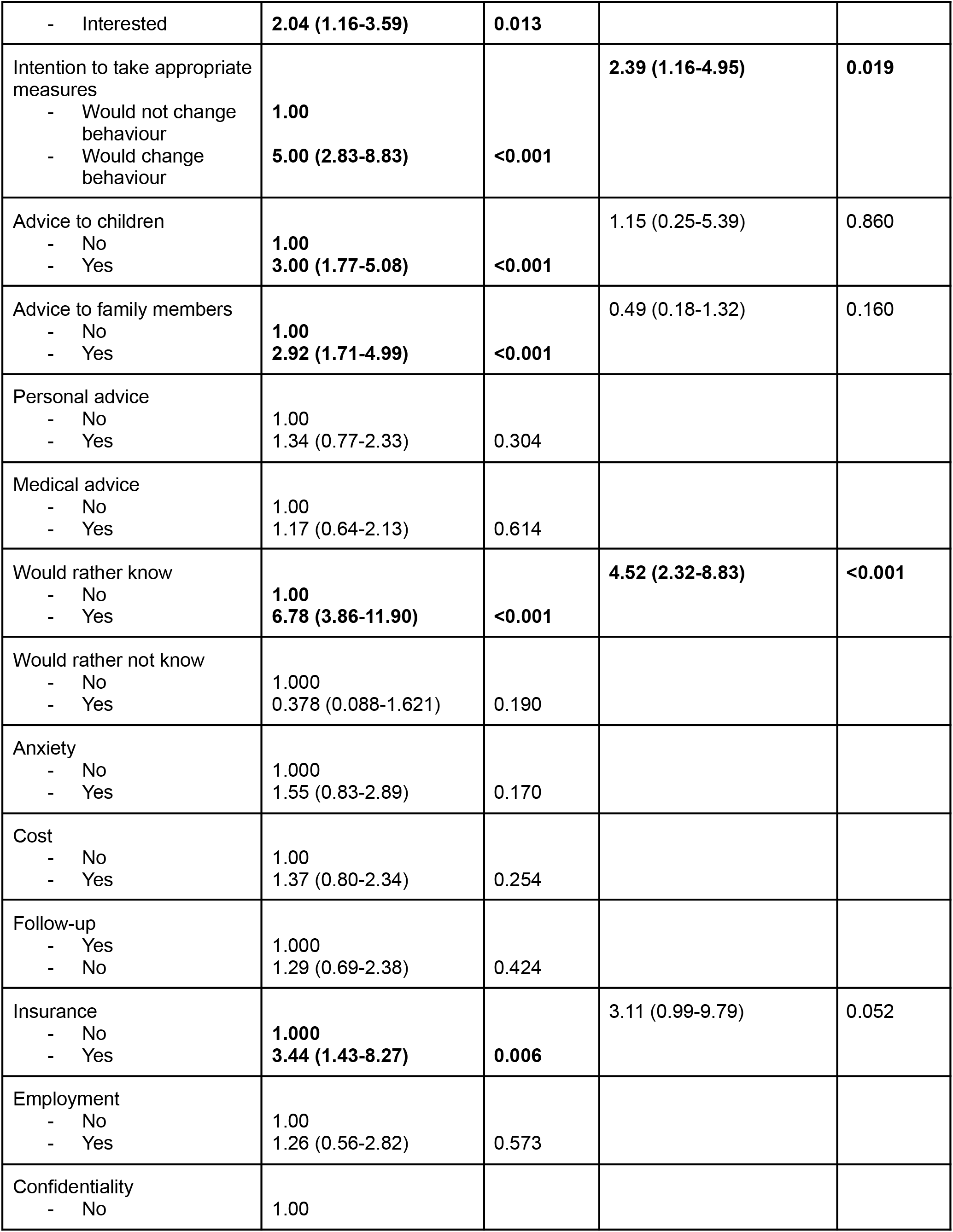
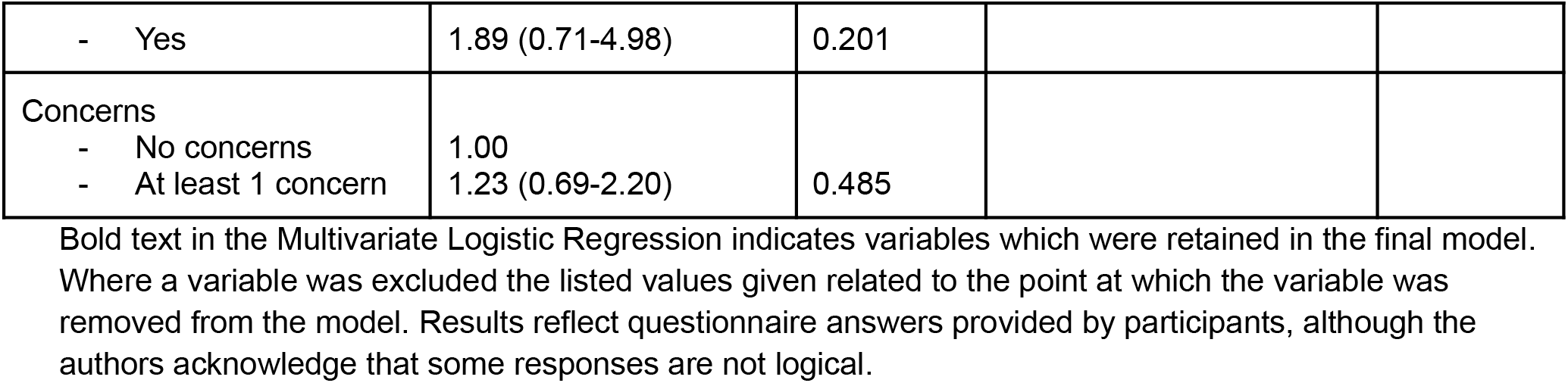
Univariate and multivariate logistic regression assessing predictors for interest in polygenic risk testing.

**Figure 1:**
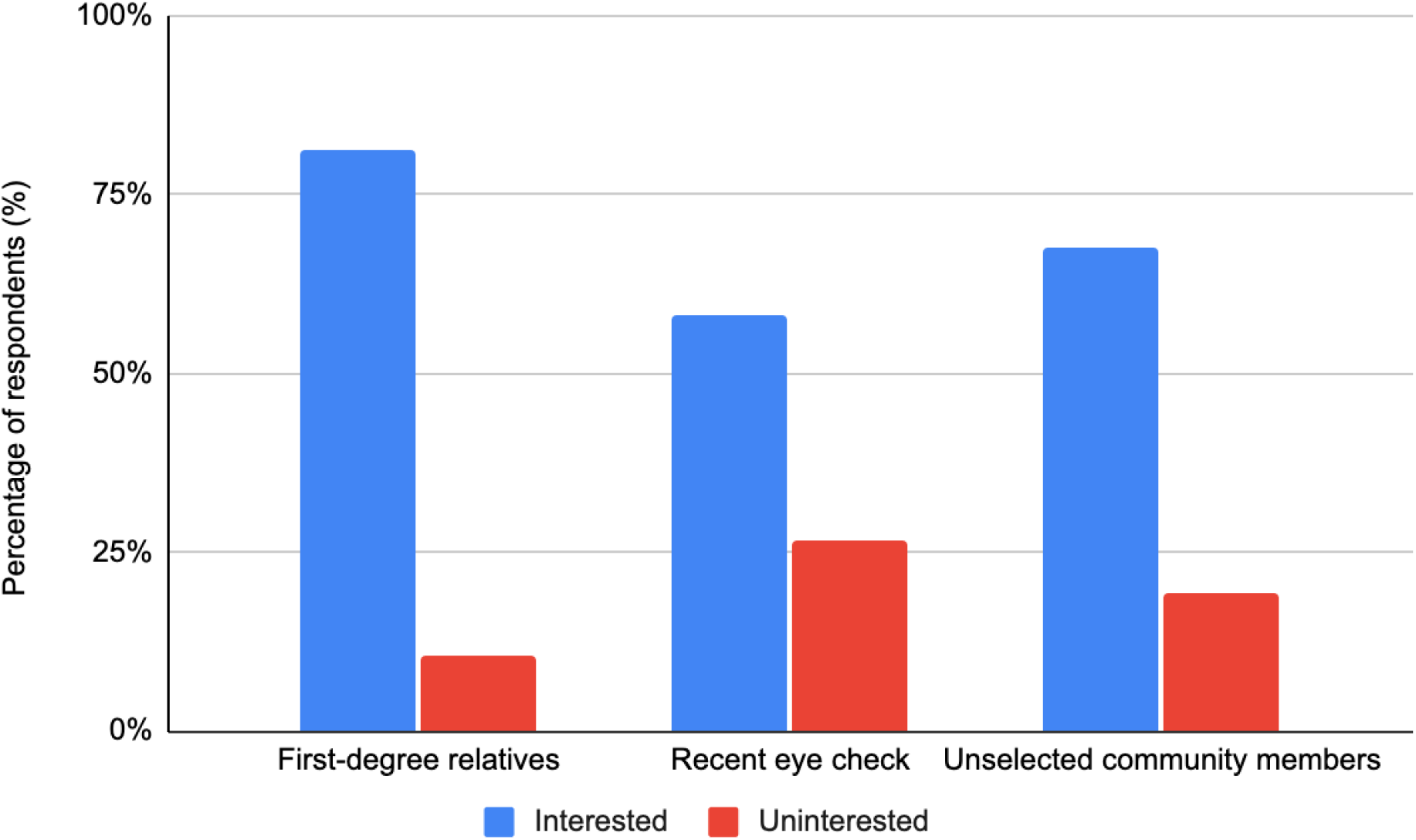
Level of interest in polygenic risk testing for glaucoma (positive versus negative) according to cohort classification. Responses to the question ‘How likely would you be to take a genetic test which could predict your risk of developing glaucoma?’. Responses were grouped by cohort classification (first-degree relatives, recent eye exam, and general members of the community), and grouped into interested (likely or highly likely) or uninterested (highly unlikely or unlikely) expressed interest. Forty-two respondents indicated being ‘unsure’ (10.0%).

### 4. Factors affecting interest in genetic risk prediction testing for glaucoma

We assessed the factors that may affect participants’ decision to be tested (Figure 2) and factors that may concern participants about genetic risk prediction testing (Figure 3). After adjusting for all variables that were significant in univariate regression, interest in glaucoma genetic risk prediction testing was more common in those who believed glaucoma to be a severe medical condition (OR 14.58, 95%CI (1.15-185.50), p = 0.039), were concerned about developing glaucoma (OR 4.37, 95%CI (2.32-8.25), p <0.001), had an intention to take appropriate measures regarding eye health (OR 2.39, 95%CI (1.16-4.95), p=0.019), or who preferred to know if they were at risk of glaucoma or not (OR 4.52, 95%CI (2.32-8.83), p<0.001) (Table 2).

**Figure 2.**
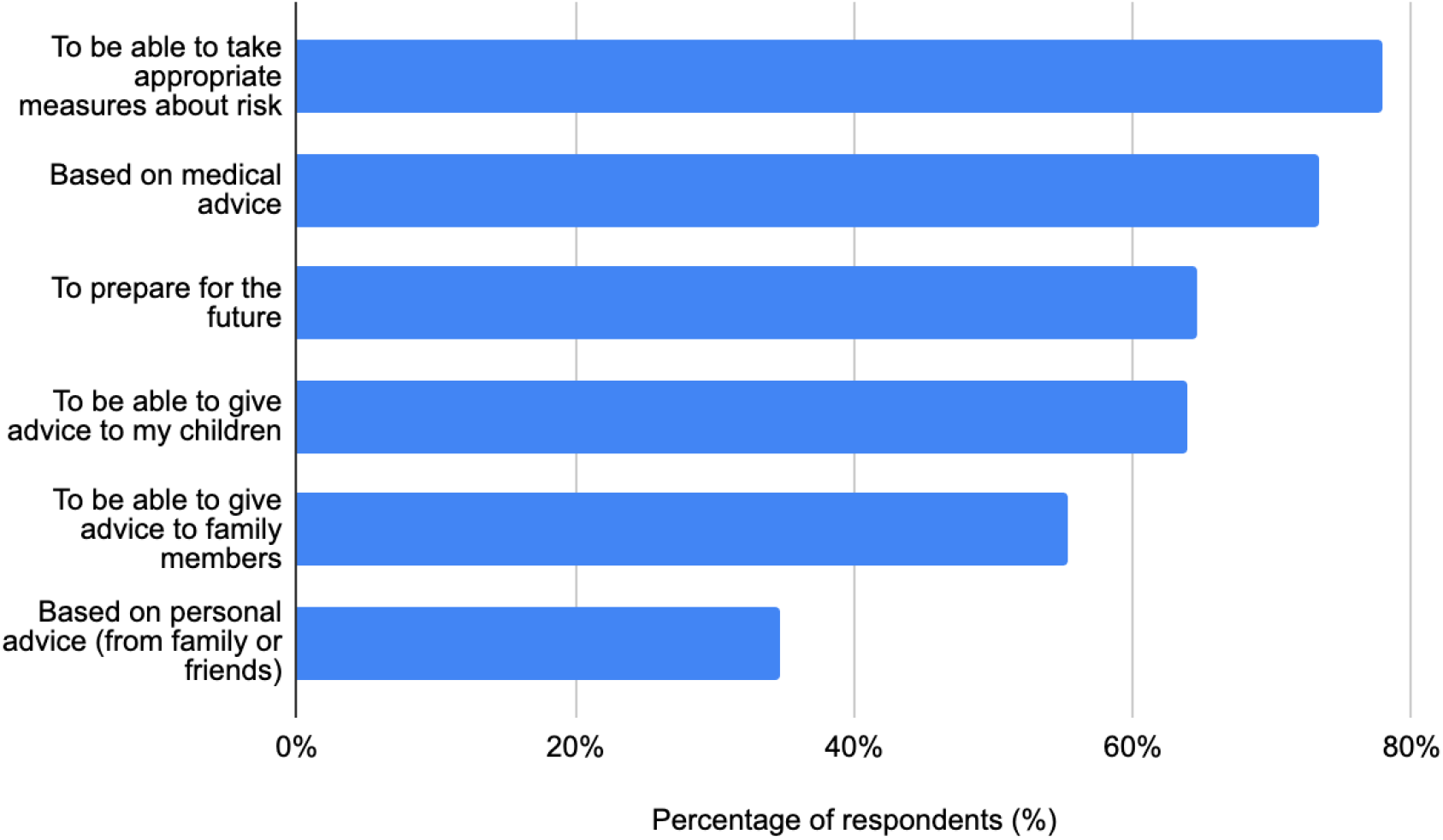
Factors affecting participants’ decision to be tested. Responses to the question ‘Which of the following factors would affect your decision to be tested? (Choose as many as appropriate)’.

**Figure 3.**
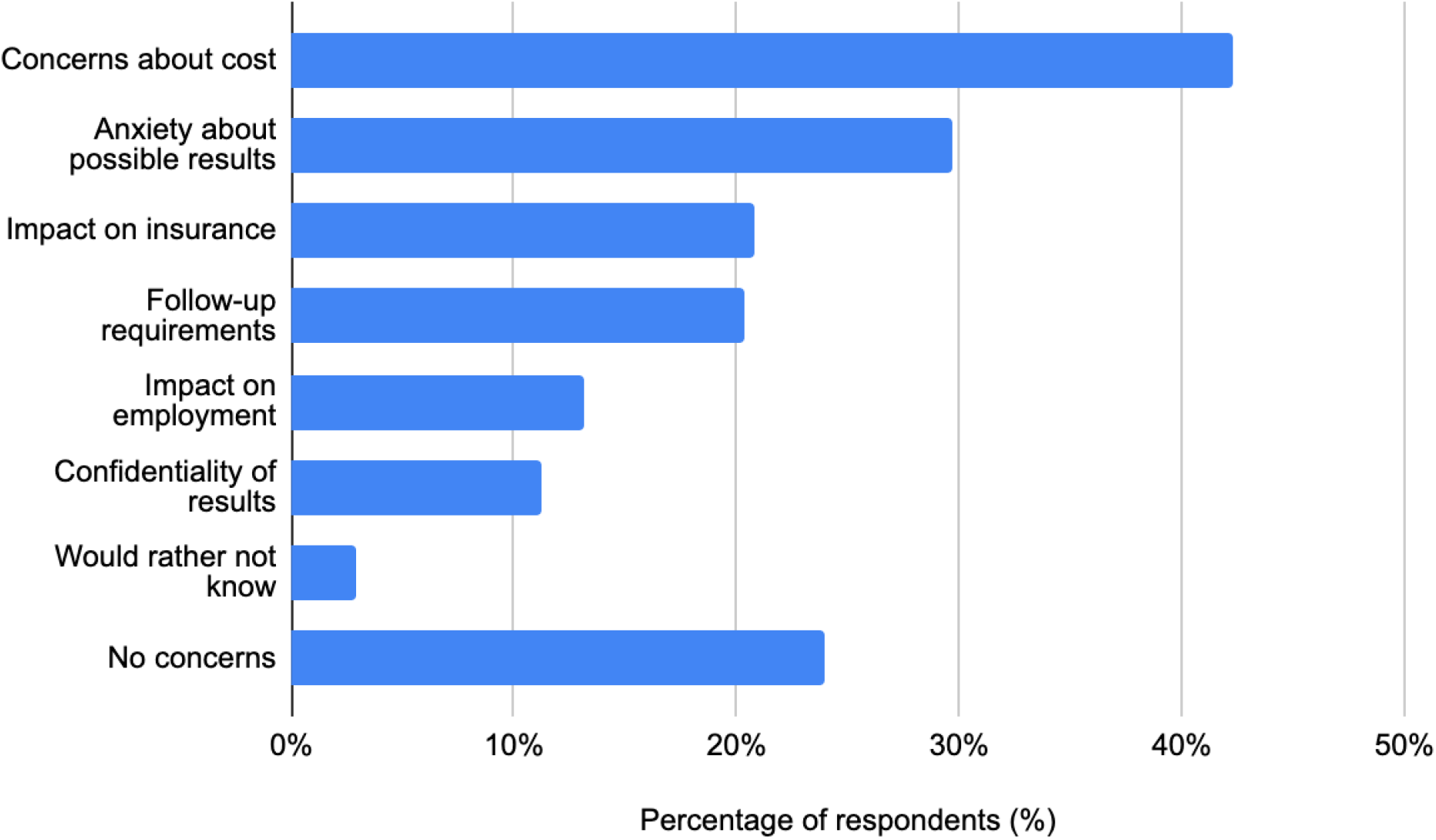
Factors concerning participants about having the test. Responses to the question ‘Which of the following factors would concern you about having the test? (Choose as many as appropriate)’.

The majority (75.8%) of individuals had at least one concern about genetic risk prediction testing for glaucoma, with cost the most frequent (42.3%), followed by personal anxiety about the possibility of the test showing increased glaucoma risk (29.7%) (Figure 3).

### 5. Behaviour

In addition to assessing which factors may influence the decision to undergo genetic risk prediction testing, we assessed whether the potential result would influence attitudes towards the frequency of future eye checks. If testing were to indicate a low risk of developing glaucoma, 91.6% of individuals indicated they would not change the frequency of their eye checks. However, if testing were to indicate a high risk of developing glaucoma, 76.6% of individuals indicated they would have more frequent eye examinations. Individuals indicated that their decision to undergo testing would be influenced more by medical advice compared to advice from family or friends (74.6% vs 35.1%, p <0.001).

### 6. Factors about testing and follow-up

Finally, we surveyed aspects of genetic risk prediction testing that participants wanted to know prior to undergoing testing. These are summarised in Supplementary Figure 1. Over 77.0% of participants deemed cost, the test process, possible implications of results, and follow-up to be important factors to understand prior to undergoing testing. Email was the most preferred method to receive results (56.5%), followed by face to face (38.3%) and letter (35.2%), with telephone call being the least preferred (21.5%). Several individuals commented that their preference would depend on the result, with face to face being preferred if results showed high glaucoma risk, and other methods, particularly email, being preferred if results showed low risk. This is consistent with the preferences indicated by individuals with glaucoma.(unpublished data) A majority of participants (64.6%) indicated they would be willing to pay at least $50 for a glaucoma genetic test if required, with AUD $50 - $100 (approximately USD $40-$70 at the time of writing) being the most acceptable range (Figure 4). Those who were willing to pay, were more likely to be interested in testing (OR 1.81, 95% CI 1.07-3.07, p = 0.028) and were also more likely to have completed tertiary education (OR 1.95, 95% CI 1.28 - 2.98, p = 0.002). Regarding the possible frequency of eye checks, approximately of all participants 88.8% indicated they would be willing to have either biannual or annual eye examinations if required (Figure 5).

**Figure 4.**
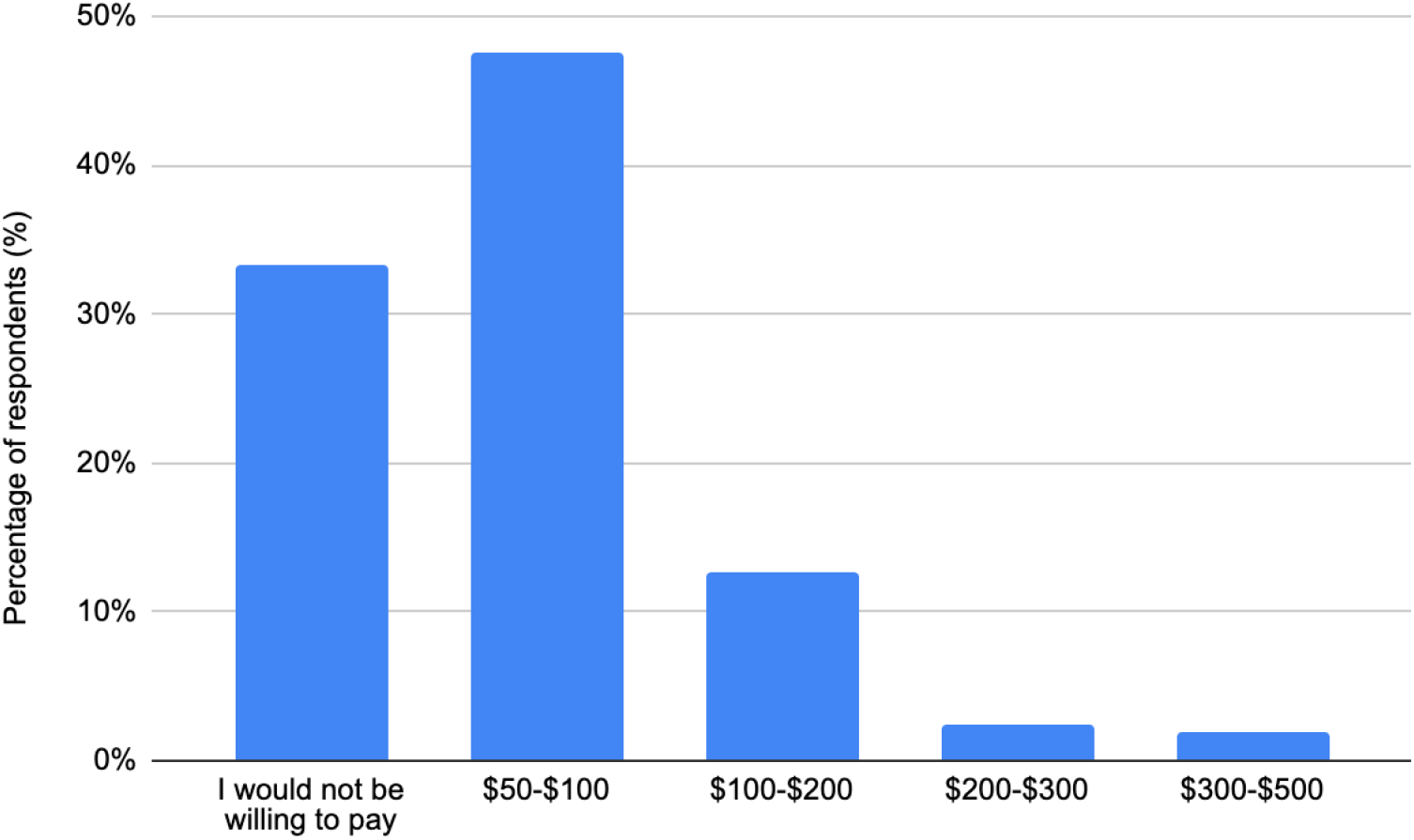
Cost participants would be willing to pay for a glaucoma genetic risk test. Responses to the question ‘If a cost were involved, how much would you be willing to pay for the test?’

**Figure 5.**
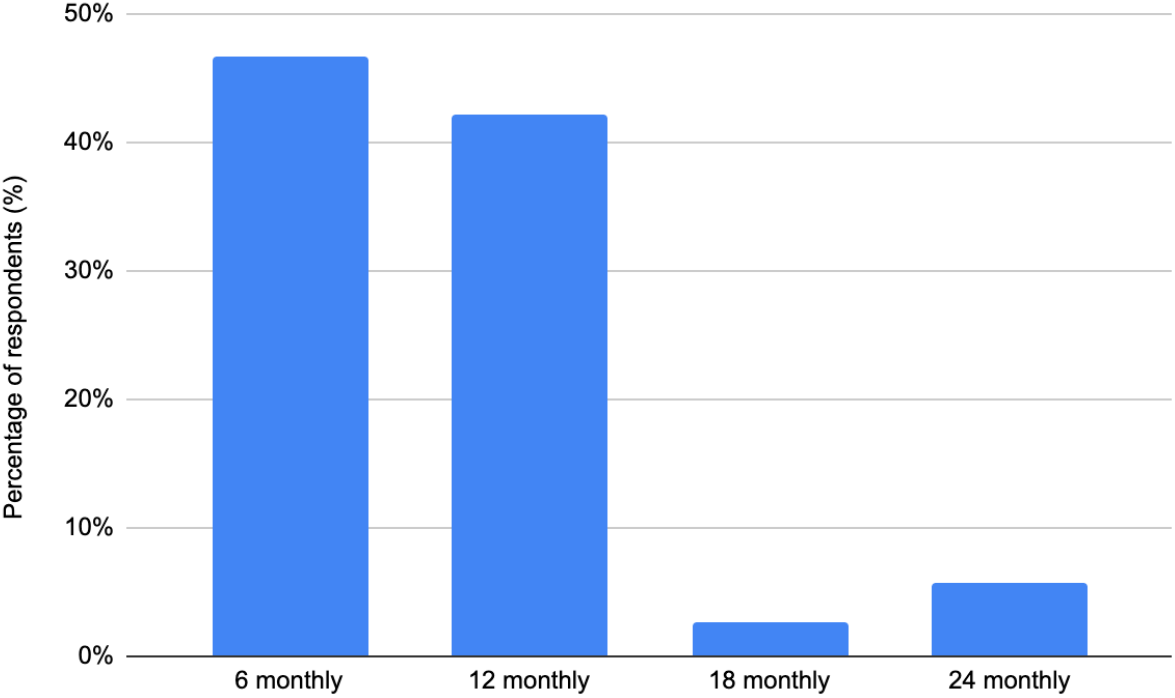
Frequency of eye checks participants would be willing to undergo. Responses to the question ‘How frequently would you be willing to have an eye check?’.

## DISCUSSION

Genetic risk stratification for diseases with complex inheritance will become increasingly accessible with the development of PRS. The majority of studies evaluating interest and attitudes toward such testing have assessed this in affected and high-risk individuals. Given one of the greatest potential advantages of PRS testing is population-scale risk stratification, it is crucial to understand the attitudes of the broader population toward this form of testing. PRS testing may be particularly useful for glaucoma given the disease’s complex heritability, lack of environmental risk factors, lack of symptoms in early disease, and success of early treatment options to slow disease progression.^22^

Our results demonstrate a similar level of interest towards PRS testing for glaucoma among unaffected individuals (71.3%) compared to individuals with diagnosed glaucoma (69.4%).(ref) Being worried about the possibility of developing glaucoma in the future appears to be a strong motivating factor to undergo testing across glaucoma and risk spectrums, with this being demonstrated in individuals with diagnosed disease and those without.(unpublished data) The attitudes of those outside of an already identified at-risk population have not been investigated for any condition. This study provides useful insights into the attitude of individuals from the community toward glaucoma genetic risk testing. Although glaucoma is the most common cause of irreversible vision loss, current screening methods are insufficient and not felt to be cost-effective at the population level.(unpublished data) Evidence of the benefit of introducing PRS testing in a broader population is growing, with previous studies showing that individuals in the top decile of a glaucoma PRS distribution reach the same absolute risk of developing the disease 10 years earlier than those in the bottom decile.^15^ This may help to guide further monitoring and treatment of high-risk individuals, as well as potentially avoiding unnecessarily regular follow-up or over-treatment of low-risk individuals.

In our study, interest in polygenic risk testing for glaucoma was associated with having a first-degree relative with the disease in the univariate logistic regression. These results are in line with several other studies which have demonstrated increased interest in PRS testing among first-degree relatives of individuals with breast cancer or colorectal cancer.^23–29^ However, while significant in a univariate logistic regression, this significance did not remain after controlling for other associated variables. This may be due to an assumed predisposition. The majority of those with an affected first-degree relative were drawn from existing glaucoma research databases. As part of their participation in these registries, individuals will have received information about the purpose of the research being to investigate the genetic nature of glaucoma, as well as additional education material about the disease. Because of this, these individuals may be aware of the risk associated with having a family history, and may therefore be undergoing regular eye examinations and feel that additional testing is not necessary. This is supported by our results which showed that those with an affected first-degree relative were not likely to change the current frequency of their eye examinations, regardless of whether a test indicated they were at either low risk (p=0.266) or high risk (p=0.073).

Those who indicated having an intention to change their behavior towards their eye health were more likely to be interested in glaucoma PRS testing. However, despite 76.7% of participants indicating being likely to have more frequent eye checks if results showed increased glaucoma risk, this was not associated with interest in PRS testing for glaucoma. This is in keeping with other studies which have shown that knowledge of risk does not correspond to a change in risk-reducing behaviours.^30,31^ Also, interest in PRS testing was associated with preferring to know their risk, rather than not. This is consistent with several studies which have shown that motivation for undergoing genetic testing commonly stems from a conviction to altruism and desire to understand more about personal health, rather than to make preventative lifestyle behaviour changes or change screening behaviours.^32–36^ The option to choose to know of a genetic susceptibility to disease may seem to be valued more than the results and their possible implications.^36^

Those who believe glaucoma to be a severe condition were more likely to be interested in PRS testing for glaucoma, and were also more likely to increase the frequency of their eye examinations if shown to be at high risk. Furthermore, those who were worried about possibly developing glaucoma in the future were also more likely to be interested in testing. This may indicate that some are motivated by fear of the possible ramifications of untreated glaucoma and potential vision loss.

Despite “having a first-degree relative with glaucoma” not reaching a statistically significant level of association with interest in testing in the multivariate logistic regression, individuals with a first-degree relative appear to recognise their risk and are worried about their potential to develop glaucoma. PRS testing may complement existing screening methods in this cohort, whereby increased risk may either be confirmed, or unnecessary concern may be alleviated in lower-risk individuals. The current National Health and Medical Research Council (NHMRC) screening guidelines in Australia recommend screening for all first-degree relatives of patients with glaucoma to commence 5 - 10 years earlier than the age of onset of glaucoma in their affected relative.^5^ Furthermore, PRS testing for glaucoma could be particularly important for those who do not have a known family history. These individuals may be unaware of any underlying risk and will not be identified early through current screening guidelines given earlier age at screening is only recommended for those with a family history.^5^

We asked participants which components of the test they would like to know more about prior to undergoing the test. The cost of the test, process involved in taking the test, implications of the results, and likely follow-up were each roughly of equal importance to respondents with over 75% indicating they would want to know. Respondents indicated email as the preferred method of receiving results, with face to face, letter and telephone call being approximately equally preferred. The majority of those who expressed willingness to pay for the test indicated $50 - $100 to be an appropriate cost for the test. While early indications of the likely cost of PRS testing are above $100, public preference is relevant in order to consider future cost subsidisation and possible impact on uptake of the test. This may reflect the study population, with many being recruited from public hospitals where the provision of health services are not associated with any out-of-pocket costs for patients. Furthermore, Medicare (Australia’s universal health insurance system) subsidises the cost of most pathology tests, thus the Australian population are generally not accustomed to paying for such tests.

Results should be interpreted in light of the study’s strengths and limitations. Of the total participants, 34.4% were drawn from existing glaucoma research registries (ANZRAG and TARRGET). These participants have previously demonstrated interest in glaucoma research, particularly regarding genetic studies and family history, and are therefore more likely to report interest in glaucoma genetic testing. However, the interest toward PRS testing was still strong among individuals who were not part of existing research projects (65.6%). 95% of our study sample was of self-reported European ancestry, highlighting the need for further validation across other ancestral backgrounds prior to implementation, which is also pertinent to the predominantly European-derived PRS instruments themselves. Furthermore, the attitudes of individuals of European ancestry may vary depending on cultural and geographic differences, such as between individuals in Australia, Northern America and Europe.

Further work needs to be done to develop an effective strategy of implementing glaucoma PRS into clinical practice to identify high-risk individuals, given whole of population glaucoma screening is currently not cost-effective. It will be important to develop cost-balanced public health policy and infrastructure which targets screening of an appropriate population to capture those at increased risk, while ensuring adequate access to screening, treatment and follow-up.^37^ Financial implications appear particularly important to individuals as it was shown that cost was the most common concern about testing and the majority indicating being unwilling to pay for testing or the minimum cost level. Further research will need to evaluate the attitude of clinicians toward PRS testing. Clinical implementation of PRS will rely on sound clinician understanding of PRS and interpretation of the significance of results, together with general acceptance of genetic risk prediction testing. Given the potential for broad population screening, ordering PRS testing, interpreting results and communication of their significance to patients will extend beyond the clinicians directly involved in glaucoma diagnosis and management. It will therefore be important to ensure relevant resources are available to upskill all clinicians who may be involved in glaucoma PRS testing.

PRS has the potential to stratify individual risk across a broad population for many common conditions, including glaucoma. We found positive interest toward glaucoma PRS testing among three different groups of unaffected individuals from the community and have identified possible target populations for initial clinical implementation. Further research should compare the uptake of PRS testing for glaucoma in those with reported interest. Acceptability of genetic risk testing by the general population is crucial for clinical implementation of such testing to be successful.

## Data Availability

The data that support the findings of this study are available from the corresponding author upon reasonable request.

## Acknowledgements

The authors thank all participants for their contribution. This project was supported by the Australian National Health and Medical Research Council (NHMRC) Centres of Research Excellence Grant (APP1116360). Emmanuelle Souzeau was supported by an Early Career Fellowship from the Hospital Research Foundation. Jamie Craig was supported by an NHMRC Practitioner Fellowship.

## Abbreviations and Acronyms

POAG: (primary open-angle glaucoma)
IOP: (intra-ocular pressure)
ANZRAG: (Australian and New Zealand Registry of Advanced Glaucoma)
(TARRGET): Targeting At Risk Relatives of Glaucoma patients for Early diagnosis and Treatment
GWAS: (genome wide association studies)

---

**Supplementary Table 1.**
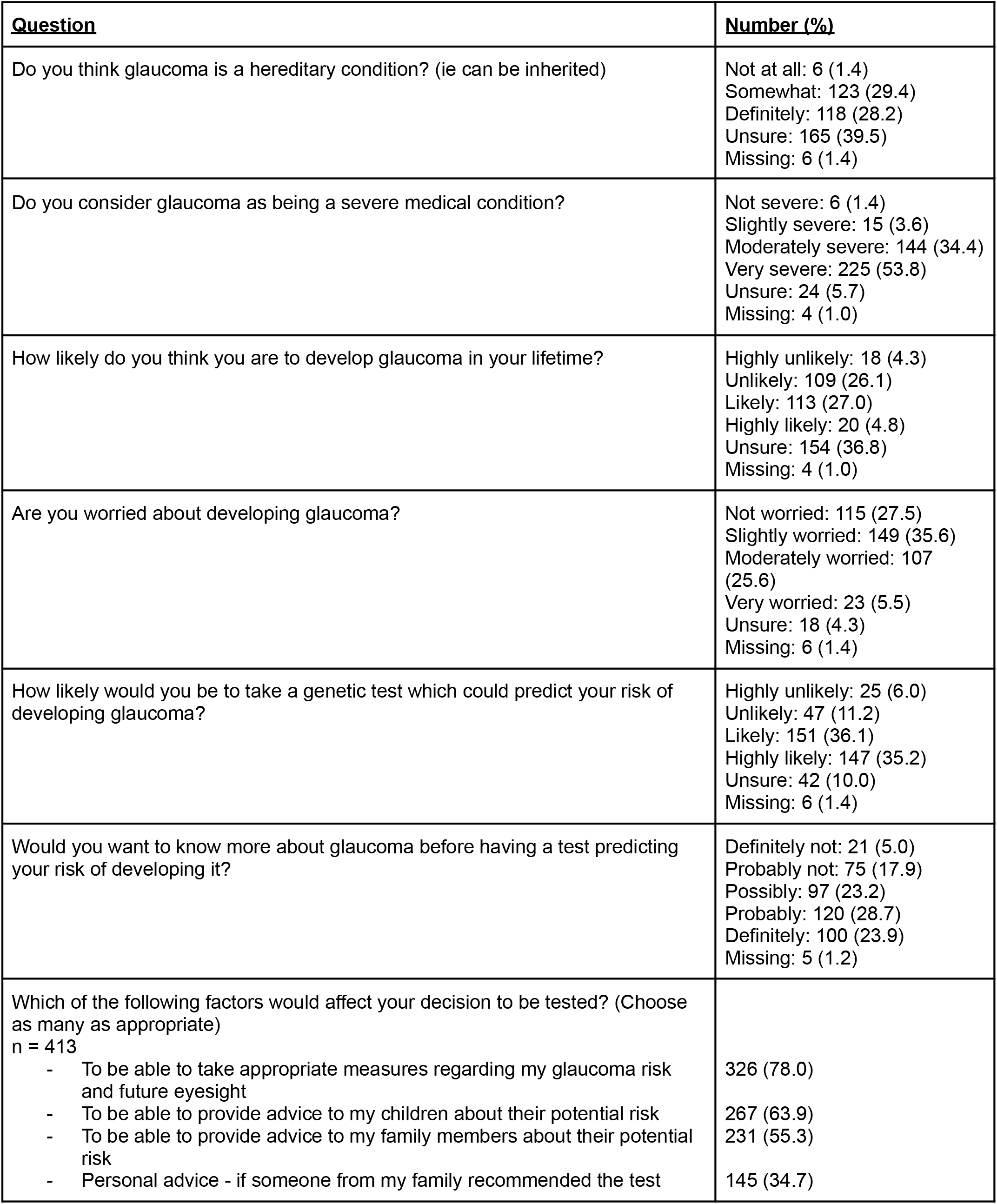
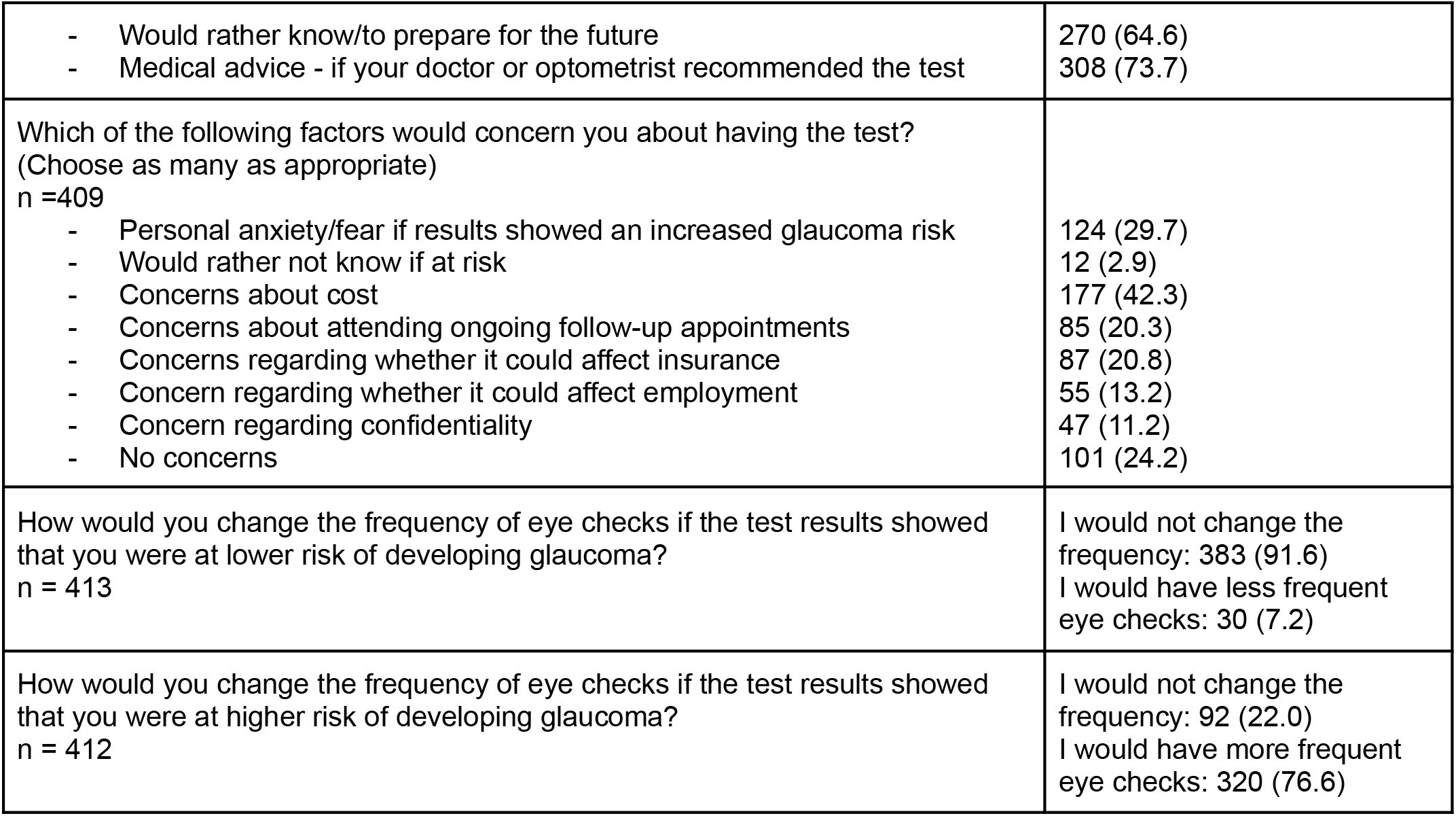
Summary of responses to survey questions relating to glaucoma and interest in testing.

**Supplementary Figure 1.**
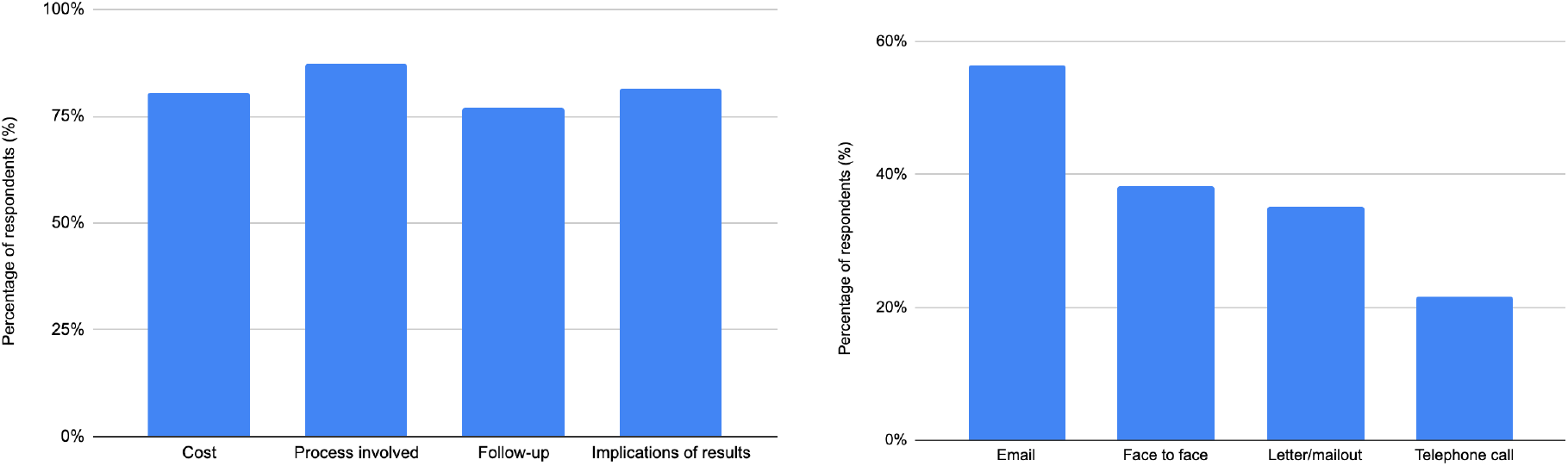
Participant’s preferences for information content surrounding genetic risk testing. Preferences were expressed for information delivered prior to having the test (A), and their preferred method of receiving the results (B). Responses to the questions ‘What information about the test would you want to know? (Choose as many as appropriate)’ and ‘What would your preferred method of receiving results be? (Choose as many as appropriate)’.

## References

1. Weinreb RN, Khaw PT. Primary open-angle glaucoma. Lancet. 2004;363(9422):1711–1720.

2. Wolfs RC, Klaver CC, Ramrattan RS, van Duijn CM, Hofman A, de Jong PT. Genetic risk of primary open-angle glaucoma. Population-based familial aggregation study. Arch Ophthalmol. 1998;116(12):1640–1645.

3. Quigley HA, Broman AT. The number of people with glaucoma worldwide in 2010 and 2020. Br J Ophthalmol. 2006;90(3):262–267.

4. Mitchell P, Smith W, Attebo K, Healey PR. Prevalence of open-angle glaucoma in Australia. The Blue Mountains Eye Study. Ophthalmology. 1996;103(10):1661–1669.

5. National Health and Medical Research Council. NHMRC Guidelines for the Screening, Prognosis, Diagnosis, Management and Prevention of Glaucoma. 2010:1–181. https://www.nhmrc.gov.au/about-us/publications/guidelines-screening-prognosis-diagnosis-management-and-prevention-glaucoma.

6. Hypertension Treatment Study O. The Ocular Hypertension Treatment Study: a randomized trial determines that topical ocular hypotensive medication delays or prevents the onset of primary open …. Archives of. 2002. https://jamanetwork.com/journals/jamaophthalmology/article-abstract/270953.

7. AGIS Investigators. The Advanced Glaucoma Intervention Study (AGIS): 9. Comparison of glaucoma outcomes in black and white patients within treatment groups. Am J Ophthalmol. 2001;132(3):311–320.

8. Charlesworth J, Kramer PL, Dyer T, et al. The Path to Open-Angle Glaucoma Gene Discovery: Endophenotypic Status of Intraocular Pressure, Cup-to-Disc Ratio, and Central Corneal Thickness. Invest Ophthalmol Vis Sci. 2010;5(7):3509–3514.

9. Springelkamp H, Iglesias AI, Mishra A, et al. New insights into the genetics of primary open-angle glaucoma based on meta-analyses of intraocular pressure and optic disc characteristics. Hum Mol Genet. 2017;26(2):438–453.

10. Wang R, Wiggs JL. Common and rare genetic risk factors for glaucoma. Cold Spring Harb Perspect Med. 2014;4(12):a017244.

11. Khawaja AP, Viswanathan AC. Are we ready for genetic testing for primary open-angle glaucoma? Eye. 2018;32(5):877–883.

12. Stone EM, Aldave AJ, Drack AV, et al. Recommendations for genetic testing of inherited eye diseases: report of the American Academy of Ophthalmology task force on genetic testing. Ophthalmology. 2012;119(11):2408–2410.

13. Dudbridge F. Power and predictive accuracy of polygenic risk scores. PLoS Genet. 2013;9(3):e1003348.

14. Torkamani A, Wineinger NE, Topol EJ. The personal and clinical utility of polygenic risk scores. Nat Rev Genet. 2018;19(9):581–590.

15. Craig JE, Han X, Qassim A, et al. Multitrait analysis of glaucoma identifies new risk loci and enables polygenic prediction of disease susceptibility and progression. Nat Genet. 2020;52(2):160–166.

16. Qassim A, Souzeau E, Siggs OM, et al. An Intraocular Pressure Polygenic Risk Score Stratifies Multiple Primary Open-Angle Glaucoma Parameters Including Treatment Intensity. Ophthalmology. 2020;127(7):901–907.

17. Gao XR, Huang H, Kim H. Polygenic Risk Score Is Associated With Intraocular Pressure and Improves Glaucoma Prediction in the UK Biobank Cohort. Transl Vis Sci Technol. 2019;8(2):10.

18. Zebardast N, Sekimitsu S, Wang J, et al. Characteristics of Gln368Ter Myocilin Variant and Influence of Polygenic Risk on Glaucoma Penetrance in the UK Biobank. Ophthalmology. March 2021. doi:10.1016/j.ophtha.2021.03.007

19. Siggs OM, Han X, Qassim A, et al. Association of Monogenic and Polygenic Risk With the Prevalence of Open-Angle Glaucoma. JAMA Ophthalmol. July 2021. doi:10.1001/jamaophthalmol.2021.2440

20. Palk AC, Dalvie S, de Vries J, Martin AR, Stein DJ. Potential use of clinical polygenic risk scores in psychiatry - ethical implications and communicating high polygenic risk. Philos Ethics Humanit Med. 2019;14(1):4.

21. Souzeau E, Goldberg I, Healey PR, et al. Australian and New Zealand Registry of Advanced Glaucoma: methodology and recruitment. Clin Experiment Ophthalmol. 2012;40(6):569–575.

22. Lewis CM, Vassos E. Prospects for using risk scores in polygenic medicine. Genome Med. 2017;9(1):96.

23. Ceballos RM, Newcomb PA, Beasley JM, Peterson S, Templeton A, Hunt JR. Colorectal cancer cases and relatives of cases indicate similar willingness to receive and disclose genetic information. Genet Test. 2008;12(3):415–420.

24. Anderson AE, Flores KG, Boonyasiriwat W, et al. Interest and informational preferences regarding genomic testing for modest increases in colorectal cancer risk. Public Health Genomics. 2014;17(1):48–60.

25. Culler DD, Silberg J, Vanner-Nicely L, Ware JL, Jackson-Cook C, Bodurtha J. Factors Influencing Men’s Interest in Gene Testing for Prostate Cancer Susceptibility. J Genet Couns. 2002;1(5):383–398.

26. Takeshima T, Okayama M, Ae R, Harada M, Kajii E. Influence of family history on the willingness of outpatients to undergo genetic testing for salt-sensitive hypertension: a cross-sectional study. BMJ Open. 2017;7(7):e016322.

27. Bottorff JL, Ratner PA, Balneaves LG, et al. Women’s interest in genetic testing for breast cancer risk: the influence of sociodemographics and knowledge. Cancer Epidemiol Biomarkers Prev. 2002;1(1):89–95.

28. Lerman C, Seay J, Balshem A, Audrain J. Interest in genetic testing among first-degree relatives of breast cancer patients. Am J Med Genet. 1995;57(3):385–392.

29. Yanes T, Meiser B, Kaur R, et al. Uptake of polygenic risk information among women at increased risk of breast cancer. Clin Genet. 2020;97(3):492–501.

30. Hollands GJ, French DP, Griffin SJ, et al. The impact of communicating genetic risks of disease on risk-reducing health behaviour: systematic review with meta-analysis. BMJ. 2016;352:i1102.

31. Bronner K, Mesters I, Weiss-Meilnik A, et al. Do individuals with a family history of colorectal cancer adhere to medical recommendations for the prevention of colorectal cancer? Familial Cancer. 2013;12(4):629–637. doi:10.1007/s10689-013-9627-x

32. Bruno M, Digennaro M, Tommasi S, et al. Attitude towards genetic testing for breast cancer susceptibility: a comparison of affected and unaffected women. Eur J Cancer Care. 2010;19(3):360–368.

33. Fraser L, Bramald S, Chapman C, et al. What motivates interest in attending a familial cancer genetics clinic? Fam Cancer. 2003;2(3-4):159–168.

34. Gollust SE, Gordon ES, Zayac C, et al. Motivations and perceptions of early adopters of personalized genomics: perspectives from research participants. Public Health Genomics. 2012;15(1):22–30.

35. Andrykowski MA, Munn RK, Studts JL. Interest in learning of personal genetic risk for cancer: a general population survey. Prev Med. 1996;25(5):527–536.

36. Murphy J, Scott J, Kaufman D, Geller G, LeRoy L, Hudson K. Public perspectives on informed consent for biobanking. Am J Public Health. 2009;99(12):2128–2134.

37. World Health Organisation. Geneva. WHO Recommendations on the Diagnosis of HIV Infection in Infants and Children. 2010. https://www.ncbi.nlm.nih.gov/books/NBK138555/.

